# Harmonizing the oral-brain axis: Music-induced microbiota shifts in age-related cognitive disorders and healthy aging

**DOI:** 10.1101/2025.07.29.25332408

**Authors:** Lúa Castelo-Martínez, Nour El Zahraa Mallah, Andrea Cavenaghi, Laura Navarro, Federico Martinón-Torres, Alberto Gómez-Carballa, Antonio Salas

## Abstract

Age-related cognitive disorders (ACD), including Alzheimer’s disease (AD), are associated with systemic inflammation and microbiome dysregulation. The oral–gut– brain axis has emerged as a critical pathway linking microbial communities to neurodegenerative processes. While music is known to influence neurological and emotional functions, its impact on the oral microbiome has not been previously explored. This study is the first to investigate how musical stimulation modulates the oral microbiome in individuals with ACD compared to cognitively healthy controls (HCs). Using next-generation sequencing (NGS), we analyzed buccal swab samples collected before and after a standardized music exposure, assessing microbial diversity, taxonomic composition, and functional pathways. Microbial diversity remained globally stable across timepoints, yet distinct taxonomic shifts emerged between groups. In ACD patients, five genera, *Veillonella*, *Porphyromonas*, *Parvimonas*, *Peptostreptococcaceae_[XI][G-9]*, and *Eikenella*, showed significant changes in abundance after musical exposure. In HCs, *Mycoplasma* was the only genus altered. At the species level, *Veillonella dispar* showed a notable reduction in ACD, highlighting its role as a key responder to music-induced modulation. Several of these taxa, including *Veillonella* and *Porphyromonas*, are well-established oral commensals with documented involvement in periodontal disease and associations with neurodegenerative processes, including AD-related neuroinflammation and amyloid pathology. Functional metagenomic analysis revealed shifts in microbial metabolic potential. Music exposure led to a downregulation of amino acid biosynthesis and upregulation of pathways involved in lipid metabolism, bile acid transformation, pantothenate/CoA biosynthesis, and taurine metabolism, all processes relevant to neurodegenerative disease mechanisms. These findings demonstrate, for the first time, that music can selectively modulate the oral microbiome at both the compositional and functional levels. This novel insight highlights the potential of music as a non-invasive modulator of microbiome– brain interactions, opening new avenues for therapeutic strategies targeting neurodegeneration through sensory stimulation.

**Key message:** This study provides the first evidence that musical stimulation can selectively modulate the oral microbiome in individuals with age-related cognitive disorders, revealing taxonomic and functional microbial shifts linked to neurodegenerative processes, highlighting music as a promising, non-invasive tool to influence microbiome–brain interactions.

## Introduction

Music has been a part of human culture for centuries, deeply intertwined with our emotions, thoughts, and even our health. It is not just about art or culture; a growing body of research shows that music can modulate brain activity, influence autonomic functions such as heart rate, and even affect endocrine responses and hormonal levels [1; 2; 3; 4; 5]. This unique and multifaceted relationship between music and physiological systems has sparked growing interest in the scientific and clinical communities regarding the potential use of music as a therapeutic modality. As a novel, non-pharmacological approach to improving mental and physical health, music therapy offers promising strategies to support patients suffering from stress, anxiety, pain, or in need of cognitive rehabilitation [6; 7; 8; 9; 10]. Understanding how music interacts with human biology may open new therapeutic avenues for promoting overall well-being and managing complex health challenges, particularly in aging populations.

In addition to our recent research [11; 12], numerous investigations [13; 14; 15; 16] have provided emerging evidence suggesting that musical stimuli can yield measurable benefits for patients with Alzheimer’s disease (AD) and other neurodegenerative disorders. These studies have employed cutting-edge methodologies from fields such as transcriptomics and genomics to examine the biological impact of music at the molecular level. Specifically, our most recent investigations have compared the influence of music on patients with age-related cognitive disorders (ACD) and healthy controls (HCs), analyzing changes in blood [11] and buccal swab samples [17]. These studies represent an important step toward understanding the biological mechanisms by which music may modulate physiological responses in ACD, shedding light on potential biomarkers and pathways influenced by auditory stimulation.

The buccal cavity has emerged as a valuable source of biological samples in biomedical research, primarily due to the feasibility of non-invasive collection methods, such as saliva and buccal swabs, along with lower processing costs compared to blood samples and their potential richness in biomarkers [18]. These samples contain a wide range of biomolecules, including proteins, hormones, antibodies, metabolites, and nucleic acids, which can reflect both local (oral) and systemic physiological states. Owing to this diverse molecular composition, saliva and buccal swab specimens have proven useful in studying a variety of health conditions, including neurodegenerative diseases such as Alzheimer’s and Parkinson’s [19; 20; 21; 22]. In this context, salivary diagnostics are gaining increasing attention as minimally invasive tools for early detection, disease monitoring, and evaluation of treatment efficacy.

AD is a progressive neurodegenerative disorder characterized by memory impairment, cognitive decline, neural loss, and the accumulation of extracellular amyloid-beta (Aβ) plaques and intracellular neurofibrillary tangles [23; 24]. While much of the current literature has concentrated on genetic mutations, tau protein dysfunction, and amyloid cascade hypotheses, recent attention has turned toward environmental and microbial factors that might influence disease progression. In this context, the role of the oral microbiome has gained interest [25; 26].

The metagenome sequencing approach enables researchers to investigate microbial diversity, taxonomic composition, and functional gene content across various biological niches, including the human oral cavity. This has opened new avenues in the study of human health and disease. Owing to its wide-ranging applications, metagenomics has become an essential tool in research on neurodegenerative diseases, particularly AD [25; 26; 27; 28; 29]. Disruptions in the oral microbiota, or dysbiosis, have been increasingly associated with systemic inflammation, neuroinflammatory signaling, and the accumulation of Aβ, all of which are key features of AD pathology [30; 31]. Through the direct sequencing of DNA from oral samples, metagenomic studies have revealed significant alterations in the microbial communities of AD patients. For example, using 16S ribosomal RNA sequencing, at the phylum level, some studies have identified increased microbial diversity in the oral microbiome of individuals with AD, with higher relative abundance of *Firmicutes* and decreased presence of *Bacteroidetes* [32]. These shifts suggest that microbial imbalance in the mouth could be linked to broader immunological and neurological consequences. Numerous studies have also identified species that may be associated with an increased risk of AD. A notable example is *Porphyromonas gingivalis*, a keystone periodontal pathogen found in the brain of AD patients [33; 34]. Additional research focusing on the genus level has reported altered microbial abundances in AD patients compared to healthy individuals [35; 36]. Nevertheless, results from the literature remain inconsistent, with an increasing number of articles aiming to characterize the microbiota composition and alterations in AD patients but failing to reach comparable results.

Building on previous findings, this study advances the field by investigating the potential influence of musical stimuli on the composition of the oral microbiome in patients with ACD and HCs. To this end, we analyzed microbial profiles derived from buccal swab samples collected from both groups, before and after exposure to music. By examining changes in microbial diversity, richness, and relative taxonomic abundance, our goal is to assess whether music can beneficially modulate the oral microbiome, potentially revealing novel biological pathways through which music may exert therapeutic effects in neurodegenerative conditions. To the best of our knowledge, this is the first study to explore the impact of music on the human microbiome using a metagenomic approach.

## Materials and methods

### Experimental design and sampling

We followed the same experimental procedures described in Gómez-Carballa et al. [11], conducted within the framework of the Sensogenomics project (http://sensogenomics.com). Briefly, the experimental concert took place at the Auditorio de Galicia on June 14, 2022. Participants were divided into two cohorts: the first consisted of individuals diagnosed with ACD, and the second, serving as HCs, included the patients’ caregivers. The musical repertoire lasted 50 minutes and featured short classical pieces performed by various combinations of a septet ensemble (the SANARTE musical group), composed of two violins, two violas, two cellos, and a horn, all professional musicians. The program included arrangements of *Spring* from *The Four Seasons* by Antonio Vivaldi, *Eine kleine Nachtmusik: Allegro* by Wolfgang Amadeus Mozart, *Pavane pour une infante défunte* by Maurice Ravel, *La musica notturna delle strade di Madrid* by Luigi Boccherini, *Rondo in E-flat major, K.371* by Mozart, *Andante for Horn and Piano* by Richard Strauss, and *Por una cabeza* by Carlos Gardel. To minimize bias, the repertoire was not disclosed to the audience beforehand. All participants remained seated and engaged only in passive listening, without physical activity during the concert. Buccal swab samples were collected using Oragene devices (ORE-100; DNAgenotek) at two timepoints: immediately before and after the concert (see Gómez-Carballa et al. [11] for detailed methodology). . The final sample included 15 ACD patients (ages 74–93, mean age 84; 73% female) and 9 HCs (ages 39–88, mean age 66; 66% female) (**Table 1**). Notably, all 9 HCs and 4 ACD patients had previously participated in the saliva gene expression study by Gómez-Carballa et al. [17].

**Table 1.**
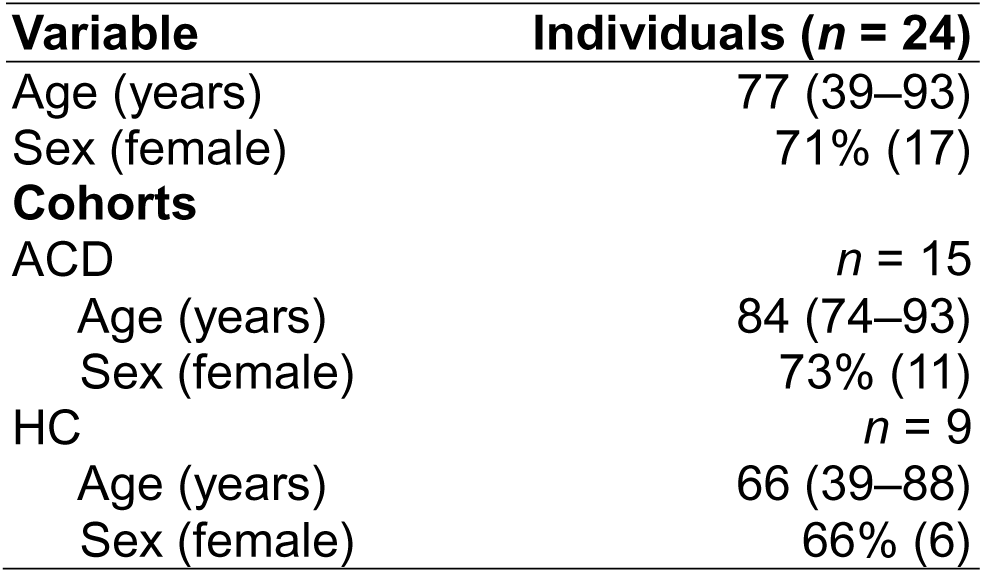
Demographic and clinical characteristics of the ACD and HC cohorts.

| **Variable** | **Individuals (*n* = 24)** |
| --- | --- |
| Age (years) | 77 (39–93) |
| Sex (female) | 71% (17) |
| **Cohorts** |  |
| ACD | *n* = 15 |
| Age (years) | 84 (74–93) |
| Sex (female) | 73% (11) |
| HC | *n* = 9 |
| Age (years) | 66 (39–88) |
| Sex (female) | 66% (6) |

All participants have permitted the publication of the project’s findings. Written informed consent was obtained from all the participants in the present study. The Ethics Committee of Xunta de Galicia approved the present project (Registration code: 2020/021), and the study was conducted in accordance with the guidelines of the Helsinki Declaration.

### DNA extraction and sequencing procedure

DNA from buccal swab samples was isolated using 500µl of sample and the *prepIT* extraction buffer (*DNAgenotek*). DNA concentration was determined using fluorometric quantification with the *Qubit* system (*Thermo Fisher Scientific Inc*.), while purity was assessed using spectrophotometry with the *NanoDrop* system (*Thermo Fisher Scientific Inc.*).

Library preparation involved amplification of the V3-V4 region of the 16S rRNA gene through two rounds of *PCR*. The resulting amplicons contained sequencing adapters compatible with *Illumina* platforms and unique indexes for sample identification. Library quality was assessed using *TapeStation* (*Agilent*) and quantified with *Qubit* fluorometry. Final libraries were quantified using *qPCR*, normalized, and pooled at equimolar concentrations to ensure optimal DNA cluster generation on the flow cell. Paired-end sequencing (2×300 bp) was performed on the *MiSeq* platform (*Illumina Inc*.), incorporating approximately 40% *PhiX* to enhance sequence diversity and maintain high-quality sequencing output. Sequencing data was generated in *FASTQ* format.

### Data processing and statistical analysis

To conduct the statistical analyses, we used R software (version 4.5.1) [37]. Quality control (QC) of the raw sequencing data was performed using the *DADA2* package (version 1.30.0) in R [38]. Initial preprocessing involved filtering and trimming to remove low-quality reads and primers, followed by error correction of the sequences. Paired-end reads (forward and reverse) were then merged to reconstruct the full denoised sequences. An amplicon sequence variant (ASV) table was generated, containing detailed information on both the samples and the inferred sequence variants. To improve the accuracy of downstream analyses, chimeric sequences were identified and removed, thereby reducing the risk of false interpretations related to microbial diversity and community composition. Finally, we assessed the number of retained reads after each pipeline step to confirm that no single process led to an excessive loss of data.

To assign the taxonomy of the sequences, we aligned the reads using the expanded *Human Oral Microbiome Database* (*eHOMD*; version 15.22 [39]) and filtered the data by prevalence and reads. Subsequently, we constructed the Operational Taxonomic Unit (OTU) table using the *Phyloseq* package (version 1.52.0) [40], focusing mainly on the relative abundance at the genus level.

To calculate the alpha (α) and beta (β) diversities, with the *MicrobiotaProcess* (version 1.14.1) [41] package, we normalized the data using the *rarefying* method by adjusting the sequence depth by the minimum library size. α*-*diversity refers to the diversity of microbial species within a single sample [42; 43], assessing the richness and evenness of microbial communities, indicating the complexity and composition of a given environment. In our case, it is described by using the *Observed OTU*, *Shannon diversity*, *Chao*’s *Richness Estimator*, and *Simpson index*. On the other hand, β-diversity measures the differences in microbial community composition between different samples, showing how microbial communities vary across environments, conditions, or individuals [44; 45]. We used *Bray-Curtis*’s dissimilarity matrix to investigate the difference between samples in taxonomic composition. Along with β-diversity, we performed a permutational multivariate analysis of variance (*PERMANOVA)* test to quantify the multivariate community-level differences between groups with the *Vegan* package (version 2.6-10) [46].

Using *MaAslin2* (version 1.22.0) [47] and *ANCOM-BC2* (version 2.10.1) packages [48], a multivariate association was performed at the genus and species levels to compare the differences between the timepoints (TP1 *vs*. TP2) and to interpret the impact of music on the oral microbiome composition, comparing both software results to derive robust information. We used a paired-samples design by including patient ID as a random effect. To ensure robustness, only taxa consistently identified as significantly different by both methods were considered for interpretation. For visual representations, the data were normalized with *Centered Log-ratio Transformation* (*CLR*) [49], and the inter-individual differences effect was removed using *ComBat* [50], with the package *MBECS* (version 1.6.0) [51], checking the distribution of the samples and the abundance of the different genera. Finally, the *ggplot* (version 3.5.1) [52] and *gg4way* (version 1.6.0) [53] packages were used to graphically represent the results.

### Functional gene prediction and pathways inference

We employed the *Tax4Fun2* R package (version 1.1.5) [54] to predict microbial functional profiles based on 16S rRNA gene (V3–V4 regions) sequencing data obtained from patients and healthy controls. Amplicon Sequence Variants (ASVs) were aligned to the NCBI RefSeq database using BLAST and mapped to corresponding 16S rRNA reference sequences. Functional inference was conducted using the pre-computed Ref100NR reference dataset, which links 16S rRNA sequences to 18,479 microbial functional profiles. Only ASVs with a sequence similarity greater than 97% were retained for functional prediction. Functional annotation was carried out using the Kyoto Encyclopedia of Genes and Genomes (KEGG) Orthology (KO) database for prokaryotes, and the predicted gene abundances were subsequently aggregated into KEGG pathways. To identify significant associations between inferred functional pathways and exposure to musical stimuli, we applied the *MaAsLin2* package (version 1.22.0) [47]. Pathway abundance data were log-transformed, and both patient ID and timepoint were modeled as random and fixed effects, respectively.

## Results

### Patterns of oral microbiota triggered by musical stimulation

To investigate the key characteristics of the oral microbiota, we first analyzed the composition and diversity of bacterial taxa in buccal samples collected from patients with a confirmed diagnosis of ACD and from HCs. Taxonomic inference was performed using the Human Oral Microbiome Database as a reference, initially identifying 2,939 taxa. A series of filtering steps was then applied to the ASV data for the ACD and HC cohorts independently. First, low-abundance taxa were excluded by retaining only those with an average read count greater than 10^-5^ across all samples. Next, the dataset was refined further by including only taxa detected more than once in at least 10% of the samples. To avoid biases in diversity estimates due to low sequencing depth, samples with fewer than 1,000 total reads were excluded. After this multi-step filtering process, 505 taxa from ACD patients and 1,045 taxa from HCs were retained for downstream analysis. Taxonomic comparisons were conducted at the genus level.

The most abundant genera in both the ACD and HC groups were *Streptococcus*, *Veillonella*, and *Prevotella* (**Figure 1A**). However, changes in relative abundance between timepoints TP1 and TP2 were more pronounced in the ACD group than in HCs. To visualize the distribution of microbiota profiles across timepoints, we performed Principal Component Analysis (PCA). PCA suggests a stronger impact of musical stimulation in ACD patients, as indicated by the variation explained by the first two principal components and the significant segregation of ACD samples along both PC1 and PC2. In contrast, HC samples exhibited separation primarily along PC2 (**Figure 1B**, **Figure S1**). These observations were further supported by heatmap analysis, which revealed distinct clustering of ACD samples by timepoint, whereas HC samples displayed only partial clustering (**Figure 1C**).

**Figure 1.**
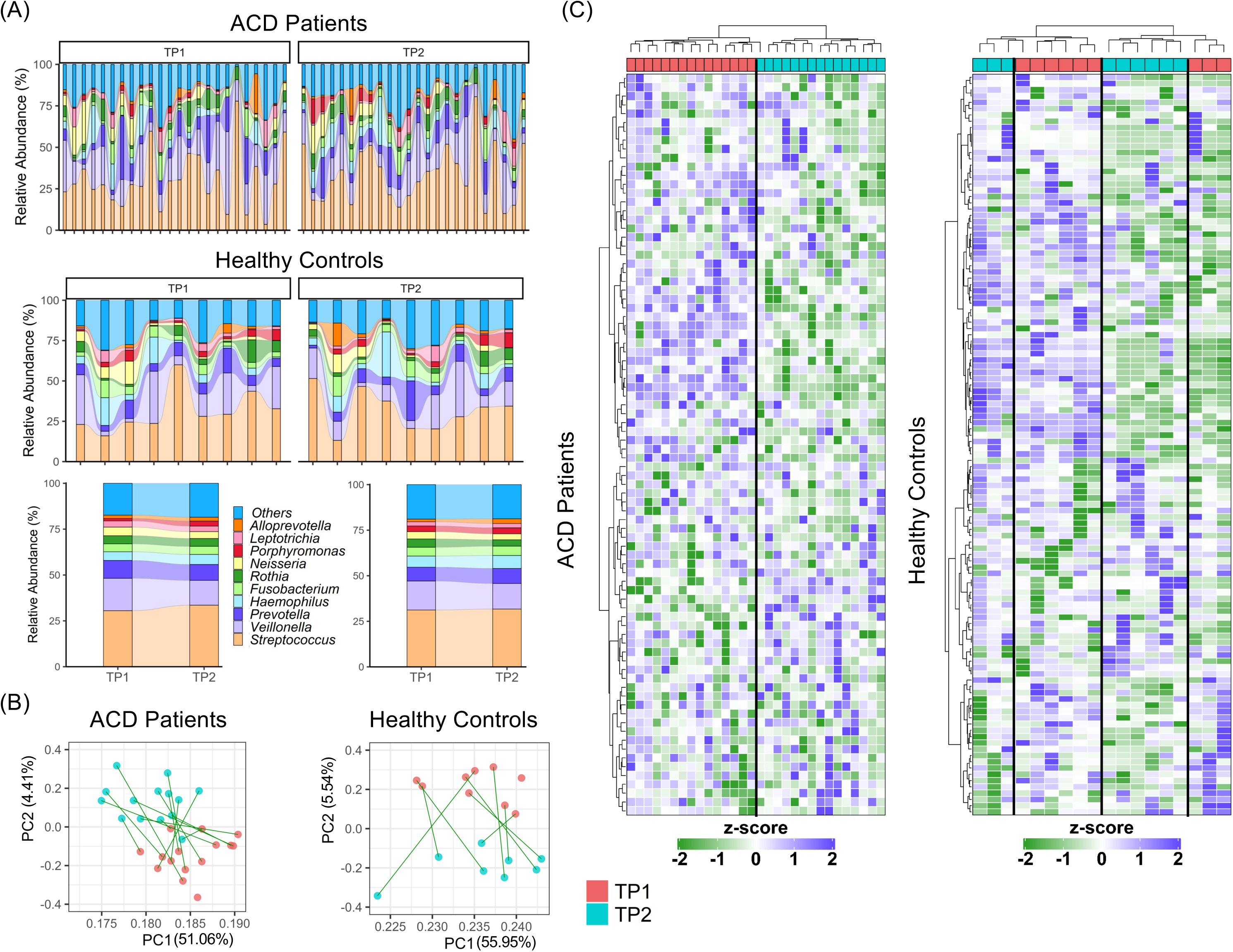
Overview of microbial community composition and structure in ACD and HC cohorts across two timepoints. A) Genus relative abundance of ACD cohort and HC cohort in the different timepoints, first by sample, followed by the abundance mean. B) PCA of ACD and HC cohorts with the distribution of the samples, connected by individual, at the two timepoints. C) Heatmap representing the abundance of all genera in the two cohorts.

To assess the influence of musical stimulation on oral microbial diversity, we calculated α-diversity (within-sample) and β-diversity (between-sample) metrics for TP1 and TP2 for both groups. α-diversity was assessed using the *Shannon*, *Chao1*, *Observed OTU*, and *Simpson* indices. We observed modest increases in median α-diversity values following the intervention in ACD patients but not in HCs, where results indicated a more heterogeneous pattern. Nevertheless, these changes were not statistically significant (*P*-value > 0.05) between TP1 and TP2 in either group (**Figure S2**).

With respect to the β-diversity, the Principal Coordinates Analysis (PCoA) based on Bray–Curtis distances did not reveal clear clustering by timepoint in either ACD or HC groups (**Figure S2**), indicating that the overall composition of the oral microbiome remained stable after the musical intervention. PERMANOVA test confirmed the absence of statistically significant differences in beta diversity between timepoints within each group (*P*-value = 0.99).

### Differential abundance analysis of oral microbiota following musical stimulation

Differential abundance analysis was first conducted at the genus level and subsequently at the species level to provide a greater resolution.

In the ACD cohort, 13 genera were identified as differentially abundant using *MaAsLin2*, and 12 genera were identified using *ANCOM-BC2* (**Figure 2A**). Notably, five genera were consistently identified by both methods: *Veillonella* (*MaAsLin2*: *P*-value = 0.003; *ANCOM-BC2*: *P*-value = 0.001), *Porphyromonas* (*MaAsLin2*: *P*-value = 0.020; *ANCOM-BC2*: *P*-value = 0.013), *Parvimonas* (*MaAsLin2*: *P*-value = 0.003; *ANCOM-BC2*: *P*-value = 0.011), *Peptostreptococcaceae_[XI][G-9]* (*MaAsLin2*: *P*-value = 0.032; *ANCOM-BC2*: *P*-value = 0.005), and *Eikenella* (*MaAsLin2*: *P*-value = 0.042; *ANCOM-BC2*: *P*-value = 0.047) (**Figure 2A**; **Tables S1** and **S2**). All of these genera, except *Veillonella*, were significantly more abundant after musical stimulation. *Veillonella* exhibited a negative Log_2_FC (Log_2_FC [*MaAsLin2*] = -0.78; Log_2_FC [*ANCOM-BC2*] = -0.71) between TP1 and TP2.

**Figure 2.**
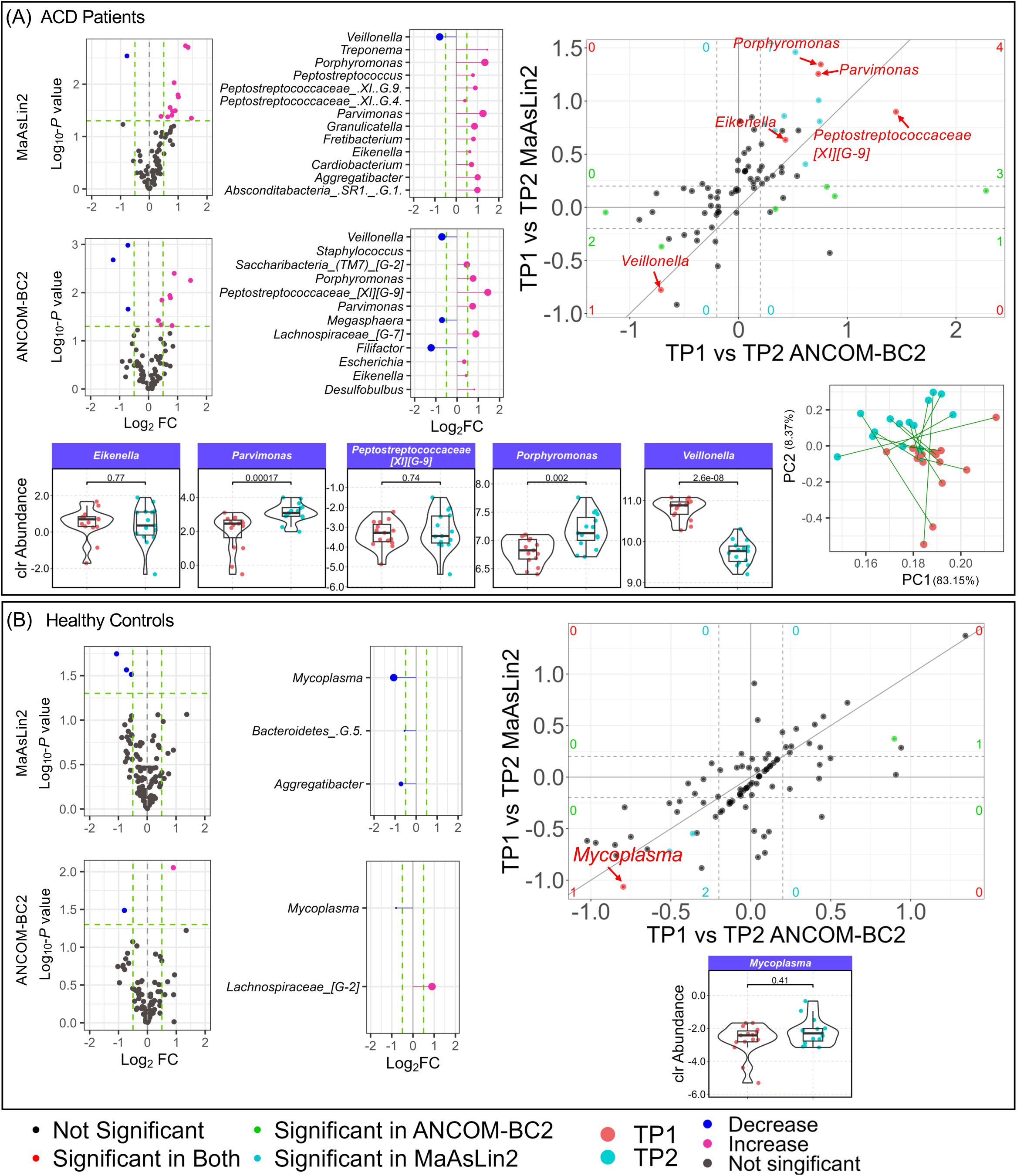
Summary of differential genus-level microbial abundance analysis in ACD and HC cohorts across timepoints. A) Differential abundance analysis results at the genus level in the ACD cohort using *MaAslin2* software and *ANCOM-BC2* software, representing the common results between the two methods, the boxplot of the corrected abundance changes in the timepoints (clr: centered log-ratio), and the PCA distribution of the samples only with the genus of interest. B) Differential abundance analysis results in HC cohort using *MaAslin2* software and *ANCOM-BC2* software, representing the common results between the two methods, and the boxplot of the corrected abundance change in the timepoints (clr: centered log-ratio).

Metagenomic profiles of these five consistently identified genera distinguished TP1 and TP2 samples in ACD patients along PC1 (accounting for 83.15% of the variation) and PC2 (8.37%), suggesting their potential sensitivity to musical intervention (**Figure 2A**). Boxplot comparisons of normalized abundances between TP1 and TP2 indicated statistically significant differences in three of the five genera: *Veillonella* (*P*-value = 2.6×10^-08^), *Parvimonas* (*P*-value = 1.7×10^-04^), and *Porphyromonas* (*P*-value = 2.0×10^-03^); **Figure 2A**.

In contrast, differential abundance analysis in the HC cohort revealed fewer changes between TP1 and TP2 (**Figure 2B**). Specifically, three genera were identified as differentially abundant using *MaAsLin2* and two using *ANCOM-BC2*, with only one overlapping genus: *Mycoplasma* (MaAsLin2: *P*-value = 0.018; *ANCOM-BC2*: *P*-value = 0.024) (**Tables S1** and **S2**). However, normalized abundance values for *Mycoplasma* did not show statistically significant differences between TP1 and TP2.

To compare overall patterns of change, we examined the correlation of results between the ACD and HC groups using both *STAT* values and Log_2_FC metrics from *MaAsLin2* and *ANCOM-BC2*. This analysis revealed no significant correlation, indicating distinct microbial response patterns to musical stimulation in the two groups (**Figure S3**).

At the species level, differential abundance analysis in the ACD group identified 14 species as significantly different between TP2 and TP1 using *MaAsLin2*, and 13 species using *ANCOM-BC2*. Only *Veillonella dispar* was identified by both methods (*MaAsLin2*: *P*-value = 0.042; *ANCOM-BC2*: *P*-value = 0.015) (**Figure 3A**; **Tables S3** and **S4**). As observed in the genus-level analysis, *Veillonella dispar* was significantly less abundant after musical intervention. In the HC cohort, three species were identified by *MaAsLin2* and 12 by *ANCOM-BC2*, with only one overlapping species showing statistically significant differences: *Saccharibacteria_(TM7)_[G-3]_bacterium_HMT_351* (*MaAsLin2*: *P*-value = 0.043; *ANCOM-BC2*: *P*-value = 0.028) (**Figure 3B**; **Tables S3** and **S4**).

**Figure 3.**
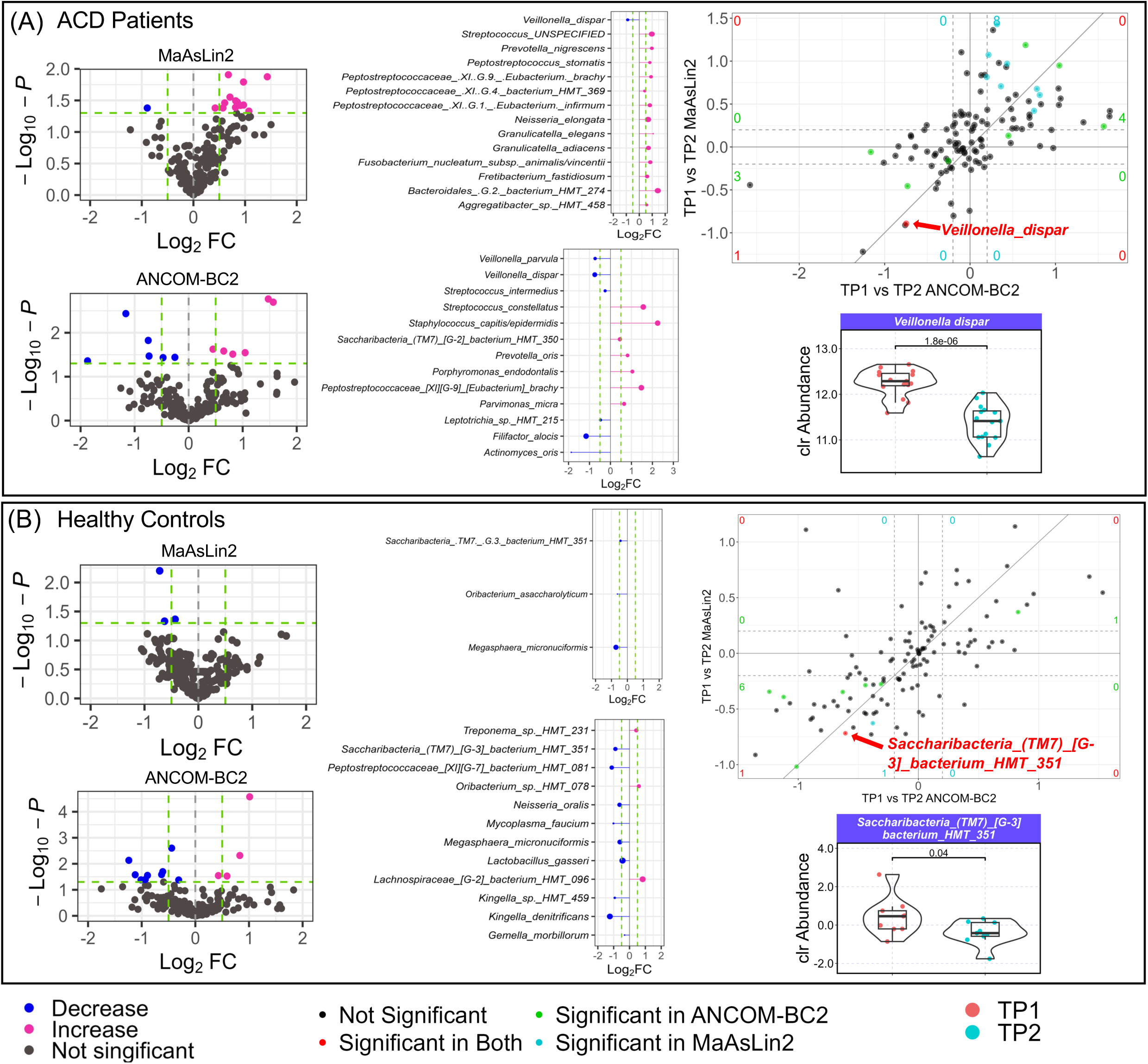
Differential abundance of microbial species across timepoints in ACD and HC cohorts. (A) Differential abundance analysis at the species level in the ACD cohort using *MaAsLin2* and *ANCOM-BC2* software, highlighting shared results between the two methods and corresponding boxplots of normalized abundance changes across timepoints (clr: centered log-ratio). (B) Differential abundance analysis in the HC cohort using *MaAsLin2* and *ANCOM-BC2* software, showing common results and boxplots of corrected abundance changes across timepoints (clr: centered log-ratio). (Note: *Streptococcus*_UNSPECIFIED represents a composite of multiple clades: *S.infantis clade 431*, *clade 638*, *S. mitis*, *S. oralis subsp. dentisani clades 058 & 398*, *S. oralis subsp. oralis*, *S. oralis subsp. tigurinus clades 070 & 071*, *S. sp. HMT 061, 064, 423*.)

### Music-induced changes in oral microbiota functions

Multivariate analysis of functional profiles revealed a total of 38 KEGG pathways significantly associated (*P*-value < 0.05; **Figure 4A**, **Table S5**) with exposure to musical stimuli (TP1 *vs*. TP2) in ACD patients, reflecting biologically relevant functional shifts in the oral microbiome. The analysis did not detect any pathway associated with the musical intervention in HCs. Most of the significantly associated pathways in ACD involved metabolic functions (26/38; 68%), followed by pathways related to human disease (7/38; 18%); **Table S5**. The analysis highlighted nine top pathways (*P*-value < 0.01; **Figure 4A**, **Table S5**) representing the most robust changes in microbial functions in response to musical stimulation.

**Figure 4.**
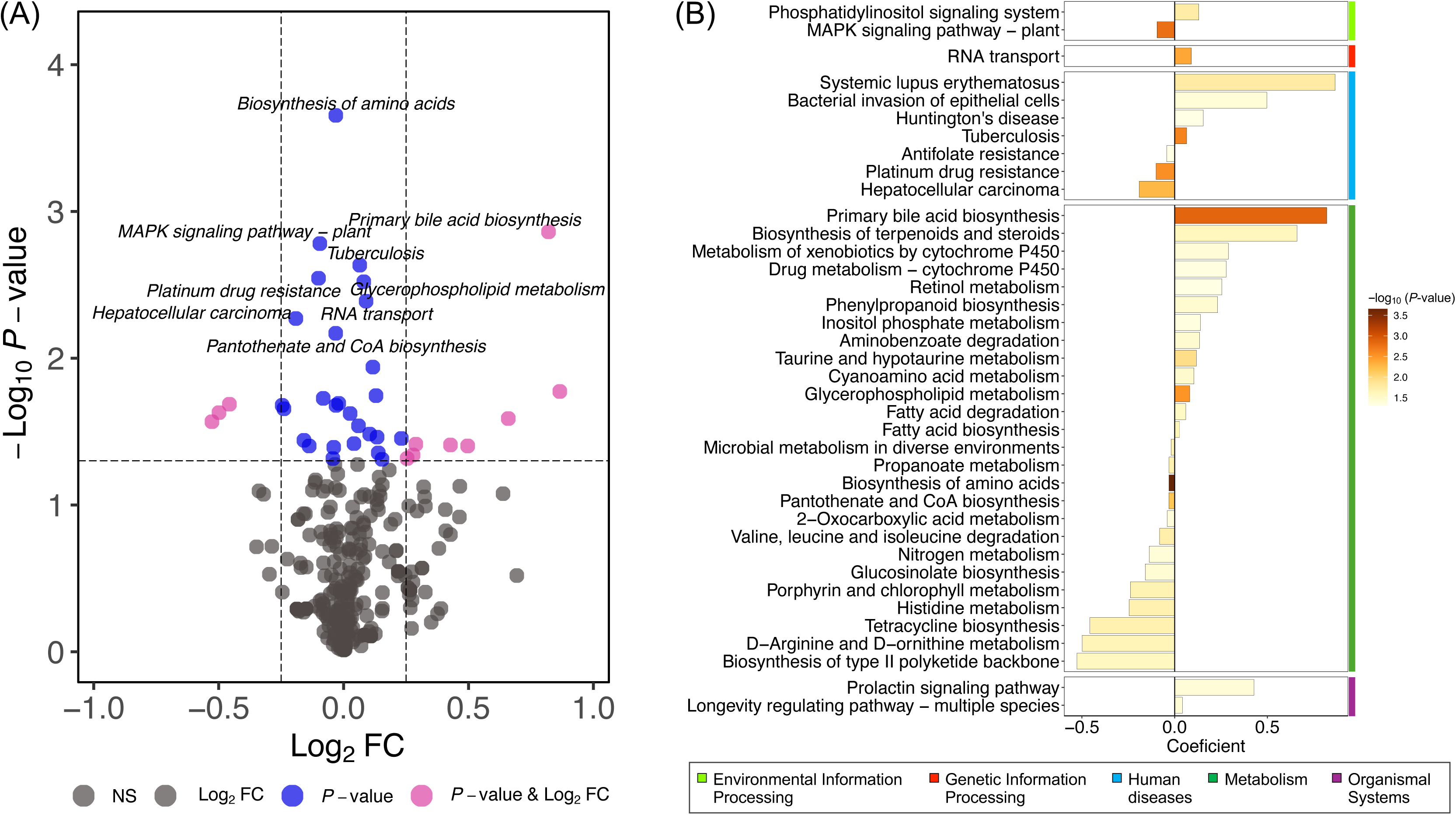
Functional pathway alterations associated with musical stimulation in the ACD cohort. (A) Volcano plot showing significant associations between inferred pathways and musical stimulation (TP1 *vs*. TP2) in ACD patients. The x-axis represents log_2_ fold change (Log_2_FC) and the y-axis shows -Log_10_ *P*-value. Pathways with *P*-value < 0.05 and |Log2FC| > 0.25 are highlighted in magenta; those with *P*-value < 0.05 and |Log2FC| < 0.25 in blue; non-significant pathways are shown in gray. Labels are provided only for pathways with *P*-value < 0.01. (B) Bar chart of significantly associated pathways (*P*-value < 0.05) in ACD patients, with bar length indicating coefficient values and color gradient representing -Log_10_ *P*-values.

The ‘biosynthesis of amino acids’ (ko01230; coefficient = –0.03, *P*-value = 2.2×10^-04^) showed the strongest association, indicating a decrease after musical exposure. Decreases were also seen in ‘valine, leucine, and isoleucine degradation’ (ko00280) and pathways involved in cofactor biosynthesis and vitamin metabolism, such as ‘pantothenate and CoA biosynthesis’ (ko00770); **Figure 4B**, **Table S5**. However, an increase in activity was detected for ‘taurine and hypotaurine metabolism’ (ko00430).

In contrast, pathways related to lipid metabolism, including ‘primary bile acid biosynthesis’, showed a significant increase in activity (ko00120; coefficient = 0.82, *P*-value = 0.0014), ‘glycerophospholipid metabolism’ (ko00564), and ‘fatty acid biosynthesis’ (ko00061), were positively associated with music exposure (**Figure 4B**, **Table S5**).

Several signaling and transport pathways were also affected. The ‘MAPK signaling pathway’ (ko04016) and the ‘phosphatidylinositol signaling system’ (ko04070) showed decreased activity after the intervention. Interestingly, enhanced microbial translational and regulatory activity was also detected through the upregulation of the ‘RNA transport’ pathway (ko03013, coefficient = 0.0901, *P*-value = 0.0041; **Figure 4B**, **Table S5**).

Changes in human disease-related functions were also observed. The ‘tuberculosis’ (ko05152) and ‘hepatocellular carcinoma’ (ko05225) pathways showed significant associations, with the latter decreasing after music exposure. A similar trend was observed for ‘platinum drug resistance’ (ko01524); **Figure 4B**, **Table S5.**

## Discussion

The scientific community increasingly recognizes the human microbiome as a critical contributor to the pathophysiology of various diseases, including ACD and, more specifically, AD [55]. Alterations in gut microbial communities have been causally linked to AD pathology, with emerging evidence pointing to hippocampal neurogenesis as a key process influenced by gut-derived and systemic factors [56]. Additionally, metabolites produced by gut microbiota are now recognized as important regulators of host metabolism and inflammation, with potential implications for both metabolic syndromes and neurodegenerative disorders [57]. As such, targeting the gut microbiome has been proposed as a promising strategy for preventing or modulating the progression of neurodegenerative diseases [58].

Building on the well-established gut–brain axis, the concept of the oral–gut–brain axis has recently emerged, incorporating the oral microbiome as an upstream component capable of influencing systemic and neurological processes [59]. Numerous studies have investigated the oral health status of individuals with AD and its association with cognitive decline. However, to date, no research has examined how music, a stimulus known to affect neurological, physiological, and emotional functions, might influence the oral microbiome, particularly in terms of microbial diversity and taxonomic composition.

Within this context, subgingival plaque represents a key microbial niche. This complex biofilm forms beneath the gumline, within the gingival sulcus, and is primarily composed of anaerobic, gram-negative bacteria strongly linked to the development of periodontal diseases such as gingivitis and periodontitis [60]. These microorganisms initiate chronic inflammatory responses, leading to gingival inflammation, connective tissue degradation, and alveolar bone loss, hallmarks of progressive periodontal pathology [61]. Given that the composition of the subgingival microbiota can differ markedly between individuals with ACD and HCs, it offers a valuable site for assessing microbiome shifts induced by musical stimulation. Investigating the differential abundance of microbial taxa in response to music may provide novel insights into how sensory interventions affect not only neurological function but also the structure and function of the oral microbiome.

Given the relevance of the oral–brain axis, which describes the bidirectional interaction between the oral cavity and the central nervous system, we explored, for the first time, the impact of musical stimuli on the oral microbiota of individuals with ACD and HCs. To this end, we replicated a previously established experimental paradigm involving salivary stimulation [17], this time applying Next Generation Sequencing (NGS) to analyze the oral microbiota at high taxonomic resolution by analyzing samples collected through buccal swabs. NGS is currently regarded as the most accurate and widely used tool in oral microbiome research, enabling comprehensive profiling of microbial communities, including uncultivable species [62; 63].

Our data show that musical stimulation did not induce significant changes in overall microbial diversity within or between samples, as evidenced by the stable α and β diversity values between the two time points. Although α-diversity showed slightly higher values in TP2 from the ACD group when compared to TP1, the results were not statistically significant in either group. Globally, this stability suggests that music-induced modulation may act through targeted microbial shifts rather than broad community restructuring. This means that, while specific taxa may vary in abundance in response to musical stimuli, the global microbial structure and diversity of the oral microbiome remain unchanged.

Certainly, clear differences in microbial abundance at the genus level were observed between TP1 and TP2 in ACD patients and HCs. Our analysis identified five genera (*Veillonella*, *Porphyromonas*, *Parvimonas*, *Peptostreptococcaceae_[XI][G-9],* and *Eikenella*) in ACD patients and one genus (*Mycoplasma*) in HCs that showed significantly different abundances after the musical intervention. Notably, the species-level analysis reinforced the genus-level results, highlighting changes in *Veillonella dispar* in ACD patients.

Many of the microorganisms affected by the musical stimuli in ACD are also known as oral commensals, but they can be involved in neurodegenerative conditions as well. As previously reported, there is an intrinsic relation between AD, oral hygiene, and periodontal disease [64]. Interventions aimed at improving oral health in patients with mild AD have been shown to potentially slow cognitive decline [65]. For instance, *Veillonella*, a gram-negative bacterium from the phylum *Firmicutes*, is typically a commensal organism found in the oral cavity. However, its increased abundance has been associated not only with the progression of periodontitis but also with systemic conditions such as AD [25; 26]. Notably, *Veillonella* was the only genus showing a significantly decreased abundance after musical stimulation and appeared to be the main contributor to reshaping the oral metagenomic composition in response to music of ACD patients. This microorganism is also notable for its ability to produce volatile sulfur compounds that contribute to oral malodor by the aid of its oral species *Veillonella dispar* and *Veillonella parvula* [66]. A longitudinal study reported that higher abundances of pro-inflammatory taxa, such as *Veillonella dispar* and *Veillonella atypica*, are associated with the progression of subjective cognitive decline [67]. In our study, species-level analysis revealed that musical intervention led to a significant reduction in *Veillonella dispar*, suggesting a potential modulatory effect of the musical stimulation. *Veillonella spp*. are also known to co-aggregate with other key periodontal pathogens such as *Fusobacterium nucleatum* and *P. gingivalis* [67]. These interactions promote biofilm formation and pathogenic synergy, thereby intensifying the inflammatory potential of the oral microbiota [68; 69]. *Porphyromonas*, *Peptostreptococcaceae_[XI][G-9]* and *Eikenella* are known contributors to periodontal infections [70; 71; 72]. Although several studies have linked cognitive decline with such infections, the exact nature of the relationship between AD and periodontitis, whether AD patients are more susceptible to periodontitis, or whether periodontitis contributes to the progression of AD remains unclear [73]. Nonetheless, *Porphyromonas* has been previously implicated as a potential factor in AD-related changes in both *in vitro* and animal studies [74; 75]. Studies have detected *P. gingivalis* in post-mortem AD brain and cerebrospinal fluid samples [76] and linked higher anti-*P. gingivalis* IgG levels in patients to cognitive impairment [77]. Experimental models showed that this microorganism or its lipopolysaccharide can induce Aβ production, pro-inflammatory responses, cognitive deficits, and reductions in neprilysin in the hippocampus [78]. Moreover, *P. gingivalis* can contribute to tau pathology, further supporting its role in AD development [74]. Another genus exhibiting differential abundance after musical stimulation in ACD, *Parvimonas*, is strongly associated with the development of periodontitis. It is known to enhance the growth of *Porphyromonas*, thereby significantly contributing to the progression of periodontal disease [79].

Inference of biological functions from metagenomic profiles indicates that musical stimuli influence primarily pathways involved in core metabolic processes. The biosynthesis of amino acids, a central metabolic route, was significantly reduced following the musical intervention, indicating a potential downregulation of general anabolic activity. Differences in levels of amino acids have been observed in AD patients [80; 81]. Genetic predisposition to elevated plasma isoleucine levels may increase the risk of developing AD [82]. However, the correlation between differences in amino acid levels and AD pathogenesis is still unclear. In AD, reduced glucose metabolism may be partially compensated by amino acid oxidation, although this shift may also promote neurotoxicity and disease progression [83]. By contrast, pathways related to lipid metabolism showed increased activity after musical stimuli, suggesting a shift in microbial capacity for lipid processing. Dysregulation of cholesterol, sphingolipids, and phospholipids has been linked to Aβ accumulation, tau hyperphosphorylation, and neuroinflammation, hallmarks in AD patients [84; 85]. In addition, increased activity in pathways involving bile acids was also identified, which are known mediators of gut-brain communication [86; 87]. Bile acids, synthesized from cholesterol in the liver and further modified by the gut microbiota. It has been hypothesized that bile acids can exert indirect neuroprotective effects in AD patients [88; 89]. Altered bile acids profiles have been detected in the serum of AD patients, likely reflecting gut microbiota dysbiosis and highlighting the potential involvement of the gut–liver–brain axis in AD pathogenesis [90]. Additionally, significant changes in pantothenate and CoA metabolism have also been observed. Pantothenate is an essential precursor for the biosynthesis of CoA. Different studies have reported global reductions of pantothenic acid levels in the brains of patients with AD and in other neurodegenerative disorders [91; 92]. Disturbances in this pathway may contribute to AD pathophysiology by impairing mitochondrial energy production, neurotransmitter synthesis, and lipid metabolism, ultimately compromising brain functions. Other significantly activated metabolic processes were related to taurine and its precursor, hypotaurine. A neuroprotective role has been attributed to taurine [93], and its abundance has been shown to decline with age [94]. This age-related reduction in taurine levels may contribute to the onset or progression of degenerative conditions. It has been reported that altered taurine metabolism, driven by gut microbiota dysregulation, could contribute to PD pathology [95]. In addition, studies in animal models underscored taurine as a potential therapeutic molecule for AD and cognitive impairment [96; 97].

A key strength of this study lies in its novelty, as it represents the first investigation into the effects of music on the oral microbiome in patients with ACD and healthy people. This innovative approach opens new avenues for exploring how non-pharmacological interventions, such as music therapy, may modulate microbiome-mediated pathways implicated in neurodegeneration.

However, several limitations should be acknowledged. The relatively small sample size may have reduced the statistical power to detect subtle microbial shifts and may limit the generalizability of the results. Additionally, the cross-sectional nature of sampling constrains our ability to infer causal relationships between musical stimulation and microbial changes.

Future studies should incorporate larger, longitudinal cohorts to validate these preliminary findings and to investigate the temporal dynamics of microbiome changes induced by musical exposure. If confirmed, music-induced microbial shifts could represent novel, non-invasive biomarkers or therapeutic targets for microbiome-modulating interventions in ACD management. Furthermore, it will be crucial to examine how factors such as oral hygiene, diet, medication use, and cognitive status interact with both musical stimulation and microbiome composition to better understand the complex interplay between lifestyle, microbial ecology, and neurodegeneration.

## Data Availability

The authors confirm that data supporting the findings of this study are available in the Sequence Read Archive (SRA) under accession numbers SRR34737793 to SRR34737812, associated with the BioProject number PRJNA1297729.

## Acknowledgements

The authors would like to express their appreciation to the study investigators of the Sensogenomics network (sensogenomics.com; Sensogenomics Working Group [see Annex]), as well as the nursery and laboratory service at the Hospital Clínico Universitario de Santiago de Compostela, for their invaluable dedication and support. This research project was made possible through the access granted by the Galician Supercomputing Center (CESGA) to its supercomputing infrastructure. The supercomputer FinisTerrae III and its permanent data storage system have been funded by the Spanish Ministry of Science and Innovation, the Galician Government, and the European Regional Development Fund (ERDF). This work was supported by: *i*) GAIN IN607B 2020/08 and IN607A 2023/02, and EUTERPE_adn (Programa de Cooperación Interreg-VI POCTEP; Ref. 0313_EUTERPE_ADN_1_E) (to A.S.), IIN607A2021/05 (to F.M.-T.) and IN677D 2024/06 (to A.G.-C.), and *ii*) Consorcio Centro de Investigación Biomédica en Red de Enfermedades Respiratorias (CB21/06/00103; to A.S. and F.M.-T.). AG-C is supported by the Miguel Servet contract (CP23/00080), funded by the Instituto de Salud Carlos III (ISCIII) and co-funded by the European Union. The funders were not involved in the study design, collection, analysis, interpretation of data, the writing of this article, or the decision to submit it for publication.

## Competing interests

The authors declare no competing interests.

## Author’s contribution

AS, FM-T, AGC, and LN conceived and coordinated the study. AS, FM-T, AGC, and LN also contributed to logistics, sample collection, and data acquisition. LCM, AGC, NZM, and AS analyzed the data and drafted the initial manuscript. AC assisted with data analysis and contributed to the interpretation and discussion of the main findings. All authors reviewed and approved the final version of the manuscript.

## Supplementary Legends

**Figure S1.** Boxplot representing the first and second dimensions of PCA in ACD and HC samples, in TP1 and TP2.

**Figure S2.** Alpha diversity (boxplots, top) and beta diversity (Principal Coordinates Analysis [PCoA] based on Bray–Curtis distances, bottom) analyses for ACD (left) and HC cohorts (right).

**Figure S3.** Correlation between the differential abundance analysis results in ACD and HC with the two methods, *MaAsLin2* and *ANCOM-BC2*, with the STAT and Log_2_FC values.

**Table S1**. Differential abundance results at the genus level using *MaAsLin2* software in ACD and HC cohorts.

**Table S2**. Differential abundance results at the genus level using *ANCOM-BC2* software in ACD and HC cohorts.

**Table S3**. Differential abundance analysis at the species level with *MaAsLin2* software comparing TP1 and TP2 in ACD and HC cohorts.

**Table S4**. Differential abundance analysis at the species level with *ANCOM-BC2* software comparing TP1 and TP2 in ACD and HC cohorts.

**Table S5**. Results from the multivariable association between *KEGG* pathways, obtained from functional gene prediction, and musical stimulation (TP1 *vs*. TP2) in ACD patients. KO: *KEGG* Orthology.

## Sensogenomics Working Group

Antonio Salas Ellacuriaga – PI; Federico Martinón-Torres – PI; Laura Navarro Ramón – Coordinator

### GenPoB/GenVip – Instituto de Investigación Sanitaria (IDIS) (alphabetical order)

Alba Camino Mera, Albert Padín Villar, Alberto G ómez Carballa, Alejandro Pérez López, Alicia Carballal Fernández, Ana Cotovad Bellas, Ana Isabel Dacosta Urbieta, Narmeen Mallah, Ana María Pastoriza Mourelle, Ana María Senín Ferreiro, Andrés Muy Pérez, Antía Rivas Oural, Antonio Justicia Grande, Antonio Piñeiro García, Anxela Cristina Delgado García, Belén Mosquera Pérez, Blanca Díaz Esteban, Carlos Durán Suárez, Carmen Curros Novo, Carmen Gómez Vieites, Carmen Rodríguez-Tenreiro Sánchez, Celia Varela Pájaro, Claudia Navarro Gonzalo, Cristina Serén Trasorras, Cristina Talavero González, Einés Monteagudo Vilavedra, Estefanía Rey Campos, Esther Montero Campos, Fernando Álvez González, Fernando Caamaño Viñas, Francisco García Iglesias, Gloria Viz Rodríguez, Hugo Alberto Tovar Velasco, Irene Álvarez Rodríguez, Irene García Zuazola, Irene Rivero Calle, Iria Afonso Carrasco, Isabel Ferreirós Vidal, Isabel Lista García, Isabel Rego Lijo, Iván Prieto Gómez, Iván Quintana Cepedal, Jacobo Pardo Seco, Jesús Eirís Puñal, José Gómez Rial, José Manuel Fernández García, José María Martinón Martínez, Julia Cela Mosquera, Julia García Currás, Julián Montoto Louzao, Lara Martínez Martínez, Laura Navarro Ramón, Lidia Piñeiro Rodríguez, Lorenzo Redondo Collazo, Lúa Castelo Martínez, Lucía Company Arciniegas, Luis Crego Rodríguez, Luisa García Vicente, Manuel Vázquez Donsión, María Dolores Martínez García, María Elena Gamborino Caramés, María Elena Sobrino Fernández, María José Currás Tuala, María Martínez Leis, María Soledad Vilas Iglesias, María Sol Rodriguez Calvo, María Teresa Autran García, Marina Casas Pérez, Marta Aldonza Torres, Marta Bouzón Alejandro, Marta Lendoiro Fuentes, Miriam Ben García, Miriam Cebey López, Montserrat López Franco, Nour El Zahraa Mallah, Narmeen Mallah, Natalia García Sánchez, Natalia Vieito Perez, Patricia Regueiro Casuso, Ricardo Suárez Camacho, Rita García Fernández, Rita Varela Estévez, Rosaura Picáns Leis, Ruth Barral Arca, Sandra Carnota Antonio, Sandra Viz Lasheras, Sara Pischedda, Sara Rey Vázquez, Sonia Marcos Alonso, Sonia Serén Fernández, Susana Rey García, Vanesa Álvarez Iglesias, Victoria Redondo Cervantes, Vanesa Álvarez Iglesias, Wiktor Dominik Nowak, Xabier Bello Paderne, Xabier Mazaira López

### Nursing volunteers (alphabetical order)

Alejandra Fernández Méndez, Ana Isabel Abadín Campaña, Ana María León Caamaño, Ana María Buide Illobre, Ángeles Mera Cores, Carmen Nieves Vastro, Carolina Suarez Crego, Concepción Rey Iglesias, Cristina Candal Regueira, Dolores Barreiro Puente, Elvira Rodríguez Rodríguez, Eugenia González Budiño, Eva Rey Álvarez, Fernando Rodríguez Gerpe, Gemma Albela Silva, Isabel Castro Pérez, Isabel Domínguez Ríos, José Ángel Fernández de la Iglesia, José Cruces Vázquez, José Luis Cambeiro Quintela, José Ramón Magariños Iglesias, Julia Rey Brandariz, Julio Abel Fernández López, Luisa García Vicente, Manuel González Lito, Manuel González Lijó, Manuela Pérez Rivas, Margarita Turnes Paredes, María Aurora Méndez López, María Begoña Tomé Arufe, María Campos Torres, María del Carmen Baloira Nogueira, María del Carmen García juan, María Esther Moricosa García, María Luz Chao Jarel, María Martínez Leis, María Mercedes Jiménez Santos, María Salomé Buide Illobre, María Victoria López Pereira, Mercedes Jorge González, Mercedes Isolina Rodríguez Rodríguez, Miren Payo Puente, Natalia Carter Domínguez, Olga María Reyes González, Pilar Mera Rodríguez, Purificación Sebio Brandariz, Salomé Quintáns lago, Yolanda Rodríguez Taboada, María Pereira Grau.

### Other volunteers (alphabetical order)

Alba Arias Gómez, Alejandro Moreno Díaz, Ana Arca Marán, Astro González Guirado, Brais García Iglesias, Carlos Sánchez Rubín, Carmen Otero de Andrés, Clara Pérez Errazquin Barrera, Claudia Rey Posse, Cristina Rojas García, Eduardo Xavier Giménez Bargiela, Elena Gloria Morales García, Fabio Izquierdo García Escribano, Gabriel Guisande García, Jaime López Martín, Lara Pais Ramiro, Lucía Rico Montero, Luís Estévez Martínez, Manuel Estévez Casal, María Aránzazu Palomino Caño, María Rubio Valdés, Marisol Nogales Benítez, Miryam Tilve Pérez, Nuria Villar Muiños, Pablo Del Cerro Rodríguez, Pablo Pozuelo Martínez Cardeñoso, Salma Ouahabi El Ouahabi, Santiago Vázquez Calvache

## Notes

### Competing Interest Statement

The authors have declared no competing interest.

### Funding Statement

This work was supported by: i) GAIN IN607B 2020/08 and IN607A 2023/02, and EUTERPE_adn (Programa de Cooperacion Interreg-VI POCTEP; Ref. 0313_EUTERPE_ADN_1_E) (to A.S.), IIN607A2021/05 (to F.M.-T.) and IN677D 2024/06 (to A.G.-C.), and ii) Consorcio Centro de Investigacion Biomedica en Red de Enfermedades Respiratorias (CB21/06/00103; to A.S. and F.M.-T.). AG-C is supported by the Miguel Servet contract (CP23/00080), funded by the Instituto de Salud Carlos III (ISCIII) and co-funded by the European Union. The funders were not involved in the study design, collection, analysis, interpretation of data, the writing of this article, or the decision to submit it for publication.

## References

[1] G.F. Halwani, P. Loui, T. Ruber, and G. Schlaug, Effects of practice and experience on the arcuate fasciculus: comparing singers, instrumentalists, and non-musicians. Front Psychol 2 (2011) 156.

[2] J. Hao, Y. Zhong, Y. Pang, Y. Jing, Y. Liu, H. Li, J. Li, and M. Zheng, The relationship between music training and cognitive flexibility: an ERP study. Front Psychol 14 (2023) 1276752.

[3] J. Kulinski, E.K. Ofori, A. Visotcky, A. Smith, R. Sparapani, and J.L. Fleg, Effects of music on the cardiovascular system. Trends Cardiovasc Med 32 (2022) 390–398.

[4] A. Linnemann, B. Ditzen, J. Strahler, J.M. Doerr, and U.M. Nater, Music listening as a means of stress reduction in daily life. Psychoneuroendocrinology 60 (2015) 82–90.

[5] Y. Song, N. Ali, and U.M. Nater, The effect of music on stress recovery. Psychoneuroendocrinology 168 (2024) 107137.

[6] E. Galinska, Music therapy in neurological rehabilitation settings. Psychiatr Pol 49 (2015) 835–46.

[7] J. Lyu, J. Zhang, H. Mu, W. Li, M. Champ, Q. Xiong, T. Gao, L. Xie, W. Jin, W. Yang, M. Cui, M. Gao, and M. Li, The Effects of Music Therapy on Cognition, Psychiatric Symptoms, and Activities of Daily Living in Patients with Alzheimer’s Disease. Journal of Alzheimer’s disease : JAD 64 (2018) 1347–1358.

[8] S.V. Moreira, F. Justi, C.F.A. Gomes, and M. Moreira, Music Therapy Enhances Episodic Memory in Alzheimer’s and Mixed Dementia: A Double-Blind Randomized Controlled Trial. Healthcare (Basel) 11 (2023).

[9] M. Sharda, C. Tuerk, R. Chowdhury, K. Jamey, N. Foster, M. Custo-Blanch, M. Tan, A. Nadig, and K. Hyde, Music improves social communication and auditory-motor connectivity in children with autism. Transl Psychiatry 8 (2018) 231.

[10] W.E. Knight, and D.N. Rickard Ph, Relaxing music prevents stress-induced increases in subjective anxiety, systolic blood pressure, and heart rate in healthy males and females. J Music Ther 38 (2001) 254–72.

[11] A. Gomez-Carballa, L. Navarro, J. Pardo-Seco, X. Bello, S. Pischedda, S. Viz-Lasheras, A. Camino-Mera, M.J. Curras, I. Ferreiros, N. Mallah, S. Rey-Vazquez, L. Redondo, A. Dacosta-Urbieta, F. Caamano-Vina, I. Rivero-Calle, C. Rodriguez-Tenreiro, F. Martinon-Torres, and A. Salas, Music compensates for altered gene expression in age-related cognitive disorders. Scientific reports 13 (2023) 21259.

[12] L. Navarro, A. Gomez-Carballa, S. Pischedda, J. Montoto-Louzao, S. Viz-Lasheras, A. Camino-Mera, T. Hinault, F. Martinon-Torres, and A. Salas, Sensogenomics of music and Alzheimer’s disease: An interdisciplinary view from neuroscience, transcriptomics, and epigenomics. Front Aging Neurosci 15 (2023) 1063536.

[13] A. Van de Winckel, H. Feys, W. De Weerdt, and R. Dom, Cognitive and behavioural effects of music-based exercises in patients with dementia. Clin Rehabil 18 (2004) 253–60.

[14] T. Särkämo, M. Tervaniemi, S. Laitinen, A. Forsblom, S. Soinila, M. Mikkonen, T. Autti, H.M. Silvennoinen, J. Erkkila, M. Laine, I. Peretz, and M. Hietanen, Music listening enhances cognitive recovery and mood after middle cerebral artery stroke. Brain : a journal of neurology 131 (2008) 866–76.

[15] A. Sisti, R. Gutman, V. Mor, L. Dionne, J.L. Rudolph, R.R. Baier, and E.M. McCreedy, Using Structured Observations to Evaluate the Effects of a Personalized Music Intervention on Agitated Behaviors and Mood in Nursing Home Residents With Dementia: Results From an Embedded, Pragmatic Randomized Controlled Trial. Am J Geriatr Psychiatry 32 (2024) 300–311.

[16] D. Kim, The Effects of a Recollection-Based Occupational Therapy Program of Alzheimer’s Disease: A Randomized Controlled Trial. Occup Ther Int 2020 (2020) 6305727.

[17] A. Gómez-Carballa, L. Navarro, N.E.-Z. Mallah, X. Bello, S. Pischedda, S. Viz-Lasheras, M.J. Currás, I. Ferreirós-Vidal, N. Mallah, J. Montoto-Louzao, A. Camino-Mera, L. Castelo-Martínez, A. Rey-Vázquez, L. Redondo, A. Dacosta-Urbieta, I. Rivero-Calle, C. Rodriguez-Tenreiro, F. Martinón-Torres, A. Salas, and S.W. Group, Music elicits different gene expression responses in the buccal cavity of age-related cognitive disorders patients and healthy controls. BioRxiv (2024) 10.1101/2024.05.29.596389.

[18] S. Chiappin, G. Antonelli, R. Gatti, and E.F. De Palo, Saliva specimen: a new laboratory tool for diagnostic and basic investigation. Clin Chim Acta 383 (2007) 30–40.

[19] K. Ngamchuea, K. Chaisiwamongkhol, C. Batchelor-McAuley, and R.G. Compton, Chemical analysis in saliva and the search for salivary biomarkers - a tutorial review. Analyst 143 (2017) 81–99.

[20] R. Farah, H. Haraty, Z. Salame, Y. Fares, D.M. Ojcius, and N. Said Sadier, Salivary biomarkers for the diagnosis and monitoring of neurological diseases. Biomed J 41 (2018) 63–87.

[21] G. Schepici, S. Silvestro, O. Trubiani, P. Bramanti, and E. Mazzon, Salivary Biomarkers: Future Approaches for Early Diagnosis of Neurodegenerative Diseases. Brain Sci 10 (2020).

[22] P. Pawlik, and K. Blochowiak, The Role of Salivary Biomarkers in the Early Diagnosis of Alzheimer’s Disease and Parkinson’s Disease. Diagnostics (Basel) 11 (2021).

[23] M.A. DeTure, and D.W. Dickson, The neuropathological diagnosis of Alzheimer’s disease. Mol Neurodegener 14 (2019) 32.

[24] R. Tenchov, J.M. Sasso, and Q.A. Zhou, Alzheimer’s Disease: Exploring the Landscape of Cognitive Decline. ACS Chem Neurosci 15 (2024) 3800–3827.

[25] D. Cammann, Y. Lu, M.J. Cummings, M.L. Zhang, J.M. Cue, J. Do, J. Ebersole, X. Chen, E.C. Oh, J.L. Cummings, and J. Chen, Genetic correlations between Alzheimer’s disease and gut microbiome genera. Scientific reports 13 (2023) 5258.

[26] H. Guo, B. Li, H. Yao, D. Liu, R. Chen, S. Zhou, Y. Ji, L. Zeng, and M. Du, Profiling the oral microbiomes in patients with Alzheimer’s disease. Oral Dis 29 (2023) 1341–1355.

[27] A.E. Perez-Cobas, L. Gomez-Valero, and C. Buchrieser, Metagenomic approaches in microbial ecology: an update on whole-genome and marker gene sequencing analyses. Microb Genom 6 (2020).

[28] A. Kaiyrlykyzy, S. Kozhakhmetov, D. Babenko, G. Zholdasbekova, D. Alzhanova, F. Olzhayev, A. Baibulatova, A.R. Kushugulova, and S. Askarova, Study of gut microbiota alterations in Alzheimer’s dementia patients from Kazakhstan. Scientific reports 12 (2022) 15115.

[29] N. Sritana, and A. Phungpinij, Analysis of Oral Microbiota in Elderly Thai Patients with Alzheimer’s Disease and Mild Cognitive Impairment. Int J Environ Res Public Health 21 (2024).

[30] A. Loughman, C.J. Adler, and H. Macpherson, Unlocking Modifiable Risk Factors for Alzheimer’s Disease: Does the Oral Microbiome Hold Some of the Keys? Journal of Alzheimer’s disease : JAD 92 (2023) 1111–1129.

[31] J. Wan, and H. Fan, Oral Microbiome and Alzheimer’s Disease. Microorganisms 11 (2023).

[32] A. Issilbayeva, A. Kaiyrlykyzy, E. Vinogradova, Z. Jarmukhanov, S. Kozhakhmetov, A. Kassenova, M. Nurgaziyev, N. Mukhanbetzhanov, D. Alzhanova, G. Zholdasbekova, S. Askarova, and A.R. Kushugulova, Oral Microbiome Stamp in Alzheimer’s Disease. Pathogens 13 (2024).

[33] T. Gong, Q. Chen, H. Mao, Y. Zhang, H. Ren, M. Xu, H. Chen, and D. Yang, Outer membrane vesicles of Porphyromonas gingivalis trigger NLRP3 inflammasome and induce neuroinflammation, tau phosphorylation, and memory dysfunction in mice. Front Cell Infect Microbiol 12 (2022) 925435.

[34] X. Ma, Y.J. Shin, J.W. Yoo, H.S. Park, and D.H. Kim, Extracellular vesicles derived from Porphyromonas gingivalis induce trigeminal nerve-mediated cognitive impairment. J Adv Res 54 (2023) 293–303.

[35] M. Taati Moghadam, N. Amirmozafari, A. Mojtahedi, B. Bakhshayesh, A. Shariati, and F. Masjedian Jazi, Association of perturbation of oral bacterial with incident of Alzheimer’s disease: A pilot study. J Clin Lab Anal 36 (2022) e24483.

[36] A. Troci, S. Philippen, P. Rausch, J. Rave, G. Weyland, K. Niemann, K. Jessen, L.P. Schmill, S. Aludin, A. Franke, D. Berg, C. Bang, and T. Bartsch, Disease- and stage-specific alterations of the oral and fecal microbiota in Alzheimer’s disease. PNAS Nexus 3 (2024) pgad427.

[37] T.R.C. Team, R: A language and environment for statistical computing. in: R.C.T. The, (Ed.), 2012.

[38] B.J. Callahan, P.J. McMurdie, M.J. Rosen, A.W. Han, A.J. Johnson, and S.P. Holmes, DADA2: High-resolution sample inference from Illumina amplicon data. Nature methods 13 (2016) 581–3.

[39] T. Chen, W.H. Yu, J. Izard, O.V. Baranova, A. Lakshmanan, and F.E. Dewhirst, The Human Oral Microbiome Database: a web accessible resource for investigating oral microbe taxonomic and genomic information. Database (Oxford) 2010 (2010) baq013.

[40] P.J. McMurdie, and S. Holmes, phyloseq: an R package for reproducible interactive analysis and graphics of microbiome census data. PLoS One 8 (2013) e61217.

[41] S. Xu, L. Zhan, W. Tang, Q. Wang, Z. Dai, L. Zhou, T. Feng, M. Chen, T. Wu, E. Hu, and G. Yu, MicrobiotaProcess: A comprehensive R package for deep mining microbiome. Innovation (Camb) 4 (2023) 100388.

[42] N.J. Gotelli, and R.K. Colwell, Quantifying biodiversity: procedures and pitfalls in the measurement and comparison of species richness. Ecol Lett 4 (2001) 379– 91.

[43] A. Brewer, and M. Williamson, A new relationship for rarefaction. Biodivers Conserv 3 (1994) 373–9.

[44] M.C. Horner-Devine, M. Lage, J.B. Hughes, and B.J. Bohannan, A taxa-area relationship for bacteria. Nature 432 (2004) 750–3.

[45] J. Jernvall, and P.C. Wright, Diversity components of impending primate extinctions. Proc Natl Acad Sci U S A 95 (1998) 11279–83.

[46] P. Dixon, VEGAN, a package of R functions for community ecology. Journal of Vegetation Science. Journal of Vegetation Science 14 (2003) 927–930.

[47] H. Mallick, A. Rahnavard, L.J. McIver, S. Ma, Y. Zhang, L.H. Nguyen, T.L. Tickle, G. Weingart, B. Ren, E.H. Schwager, S. Chatterjee, K.N. Thompson, J.E. Wilkinson, A. Subramanian, Y. Lu, L. Waldron, J.N. Paulson, E.A. Franzosa, H.C. Bravo, and C. Huttenhower, Multivariable association discovery in population-scale meta-omics studies. PLoS computational biology 17 (2021) e1009442.

[48] H. Lin, and S.D. Peddada, Multigroup analysis of compositions of microbiomes with covariate adjustments and repeated measures. Nature methods 21 (2024) 83–91.

[49] M. Kucera, and B.A. Malmgren, Logratio transformation of compositional data: a resolution of the constant sum constraint. Mar Micropaleontol 34 (1998) 117–120.

[50] W.E. Johnson, C. Li, and A. Rabinovic, Adjusting batch effects in microarray expression data using empirical Bayes methods. Biostatistics 8 (2007) 118–27.

[51] M. Olbrich, A. Kunstner, and H. Busch, MBECS: Microbiome Batch Effects Correction Suite. BMC Bioinformatics 24 (2023) 182.

[52] H. Wickham, ggplot2: Elegant Graphics for Data Analysis, Springer-Verlag New York, 2016.

[53] B.I. Laufer, and B.A. Friedman, gg4way: 4way Plots of Differential Expression. in: Bioconductor, (Ed.), 2025.

[54] F. Wemheuer, J.A. Taylor, R. Daniel, E. Johnston, P. Meinicke, T. Thomas, and B. Wemheuer, Tax4Fun2: prediction of habitat-specific functional profiles and functional redundancy based on 16S rRNA gene sequences. Environ Microbiome 15 (2020) 11.

[55] G.E. Leonov, Y.R. Varaeva, E.N. Livantsova, and A.V. Starodubova, The Complicated Relationship of Short-Chain Fatty Acids and Oral Microbiome: A Narrative Review. Biomedicines 11 (2023).

[56] S. Grabrucker, M. Marizzoni, E. Silajdzic, N. Lopizzo, E. Mombelli, S. Nicolas, S. Dohm-Hansen, C. Scassellati, D.V. Moretti, M. Rosa, K. Hoffmann, J.F. Cryan, O.F. O’Leary, J.A. English, A. Lavelle, C. O’Neill, S. Thuret, A. Cattaneo, and Y.M. Nolan, Microbiota from Alzheimer’s patients induce deficits in cognition and hippocampal neurogenesis. Brain : a journal of neurology 146 (2023) 4916–4934.

[57] Y.H. Jung, C.W. Chae, and H.J. Han, The potential role of gut microbiota-derived metabolites as regulators of metabolic syndrome-associated mitochondrial and endolysosomal dysfunction in Alzheimer’s disease. Exp Mol Med 56 (2024) 1691–1702.

[58] A. Varesi, E. Pierella, M. Romeo, G.B. Piccini, C. Alfano, G. Bjorklund, A. Oppong, G. Ricevuti, C. Esposito, S. Chirumbolo, and A. Pascale, The Potential Role of Gut Microbiota in Alzheimer’s Disease: From Diagnosis to Treatment. Nutrients 14 (2022).

[59] L.D. Sansores-Espana, S. Melgar-Rodriguez, K. Olivares-Sagredo, E.A. Cafferata, V.M. Martinez-Aguilar, R. Vernal, A.C. Paula-Lima, and J. Diaz-Zuniga, Oral-Gut-Brain Axis in Experimental Models of Periodontitis: Associating Gut Dysbiosis With Neurodegenerative Diseases. Front Aging 2 (2021) 781582.

[60] R. Doherty, Biofilms: What does subgingival plaque look like? Br Dent J 221 (2016) 16.

[61] J. Waerhaug, Subgingival plaque and loss of attachment in periodontosis as evaluated on extracted teeth. J Periodontol 48 (1977) 125–30.

[62] E.J. Stewart, Growing unculturable bacteria. Journal of bacteriology 194 (2012) 4151–60.

[63] P.E. Fournier, G. Dubourg, and D. Raoult, Clinical detection and characterization of bacterial pathogens in the genomics era. Genome Med 6 (2014) 114.

[64] A. Shahin, Y.A. Leshem, Y. Taieb, S. Baum, A. Barzilai, D. Jeddah, E. Sharon, O. Koren, R. Tzach-Nahman, S. Coppenhagen-Glazer, R. Hazan, Y. Houri-Haddad, and S. Greenberger, Association of Adult Atopic Dermatitis with Impaired Oral Health and Oral Dysbiosis: A Case-Control Study. Int Dent J 75 (2025) 279–287.

[65] L. Chen, H. Cao, X. Wu, X. Xu, X. Ji, B. Wang, P. Zhang, and H. Li, Effects of oral health intervention strategies on cognition and microbiota alterations in patients with mild Alzheimer’s disease: A randomized controlled trial. Geriatr Nurs 48 (2022) 103–110.

[66] J. Washio, Y. Shimada, M. Yamada, R. Sakamaki, and N. Takahashi, Effects of pH and lactate on hydrogen sulfide production by oral Veillonella spp. Appl Environ Microbiol 80 (2014) 4184–8.

[67] C. Ma, Y. Li, Z. Mei, C. Yuan, J.H. Kang, F. Grodstein, A. Ascherio, W.C. Willett, A.T. Chan, C. Huttenhower, M.J. Stampfer, and D.D. Wang, Association Between Bowel Movement Pattern and Cognitive Function: Prospective Cohort Study and a Metagenomic Analysis of the Gut Microbiome. Neurology 101 (2023) e2014–e2025.

[68] P. Zhou, X. Li, I.H. Huang, and F. Qi, Veillonella Catalase Protects the Growth of Fusobacterium nucleatum in Microaerophilic and Streptococcus gordonii-Resident Environments. Appl Environ Microbiol 83 (2017).

[69] A. Sakanaka, M. Kuboniwa, S. Shimma, S.A. Alghamdi, S. Mayumi, R.J. Lamont, E. Fukusaki, and A. Amano, Fusobacterium nucleatum Metabolically Integrates Commensals and Pathogens in Oral Biofilms. mSystems 7 (2022) e0017022.

[70] J. Mysak, S. Podzimek, P. Sommerova, Y. Lyuya-Mi, J. Bartova, T. Janatova, J. Prochazkova, and J. Duskova, Porphyromonas gingivalis: major periodontopathic pathogen overview. J Immunol Res 2014 (2014) 476068.

[71] C.K. Chen, and M.E. Wilson, Eikenella corrodens in human oral and non-oral infections: a review. J Periodontol 63 (1992) 941–53.

[72] G.I. Lafaurie, Y. Neuta, R. Rios, M. Pacheco-Montealegre, R. Pianeta, D.M. Castillo, D. Herrera, J. Reyes, L. Diaz, Y. Castillo, M. Sanz, and M. Iniesta, Differences in the subgingival microbiome according to stage of periodontitis: A comparison of two geographic regions. PLoS One 17 (2022) e0273523.

[73] S. Mao, C.P. Huang, H. Lan, H.G. Lau, C.P. Chiang, and Y.W. Chen, Association of periodontitis and oral microbiomes with Alzheimer’s disease: A narrative systematic review. J Dent Sci 17 (2022) 1762–1779.

[74] S.S. Dominy, C. Lynch, F. Ermini, M. Benedyk, A. Marczyk, A. Konradi, M. Nguyen, U. Haditsch, D. Raha, C. Griffin, L.J. Holsinger, S. Arastu-Kapur, S. Kaba, A. Lee, M.I. Ryder, B. Potempa, P. Mydel, A. Hellvard, K. Adamowicz, H. Hasturk, G.D. Walker, E.C. Reynolds, R.L.M. Faull, M.A. Curtis, M. Dragunow, and J. Potempa, Porphyromonas gingivalis in Alzheimer’s disease brains: Evidence for disease causation and treatment with small-molecule inhibitors. Science advances 5 (2019) eaau3333.

[75] G. Jungbauer, A. Stahli, X. Zhu, L. Auber Alberi, A. Sculean, and S. Eick, Periodontal microorganisms and Alzheimer disease - A causative relationship? Periodontol 2000 89 (2022) 59–82.

[76] C. Weber, A. Dilthey, and P. Finzer, The role of microbiome-host interactions in the development of Alzheimer s disease. Front Cell Infect Microbiol 13 (2023) 1151021.

[77] J.M. Noble, L.N. Borrell, P.N. Papapanou, M.S. Elkind, N. Scarmeas, and C.B. Wright, Periodontitis is associated with cognitive impairment among older adults: analysis of NHANES-III. Journal of neurology, neurosurgery, and psychiatry 80 (2009) 1206–11.

[78] T. Morikawa, O. Uehara, D. Paudel, K. Yoshida, F. Harada, D. Hiraki, J. Sato, H. Matsuoka, Y. Kuramitsu, M. Michikawa, and Y. Abiko, Systemic Administration of Lipopolysaccharide from Porphyromonas gingivalis Decreases Neprilysin Expression in the Mouse Hippocampus. In Vivo 37 (2023) 163–172.

[79] J. Neilands, J.R. Davies, F.J. Bikker, and G. Svensater, Parvimonas micra stimulates expression of gingipains from Porphyromonas gingivalis in multi-species communities. Anaerobe 55 (2019) 54–60.

[80] X. Ma, X.M. Wang, G.Z. Tang, Y. Wang, X.C. Liu, S.D. Wang, P. Peng, X.H. Qi, X.Y. Qin, Y.J. Wang, C.W. Wang, and J.N. Zhou, Alterations of amino acids in older adults with Alzheimer’s Disease and Vascular Dementia. Amino Acids 57 (2025) 10.

[81] H. Jesko, A. Wilkaniec, M. Cieslik, W. Hilgier, M. Gassowska, W.J. Lukiw, and A. Adamczyk, Altered Arginine Metabolism in Cells Transfected with Human Wild-Type Beta Amyloid Precursor Protein (betaAPP). Curr Alzheimer Res 13 (2016) 1030–9.

[82] S.C. Larsson, and H.S. Markus, Branched-chain amino acids and Alzheimer’s disease: a Mendelian randomization analysis. Scientific reports 7 (2017) 13604.

[83] J.W. Griffin, and P.C. Bradshaw, Amino Acid Catabolism in Alzheimer’s Disease Brain: Friend or Foe? Oxid Med Cell Longev 2017 (2017) 5472792.

[84] H. Chew, V.A. Solomon, and A.N. Fonteh, Involvement of Lipids in Alzheimer’s Disease Pathology and Potential Therapies. Frontiers in physiology 11 (2020) 598.

[85] S. He, Z. Xu, and X. Han, Lipidome disruption in Alzheimer’s disease brain: detection, pathological mechanisms, and therapeutic implications. Mol Neurodegener 20 (2025) 11.

[86] V.F. Monteiro-Cardoso, M. Corliano, and R.R. Singaraja, Bile Acids: A Communication Channel in the Gut-Brain Axis. Neuromolecular Med 23 (2021) 99–117.

[87] J.M. Ferrell, and J.Y.L. Chiang, Bile acid receptors and signaling crosstalk in the liver, gut and brain. Liver Res 5 (2021) 105–118.

[88] A. Mulak, Bile Acids as Key Modulators of the Brain-Gut-Microbiota Axis in Alzheimer’s Disease. Journal of Alzheimer’s disease : JAD 84 (2021) 461–477.

[89] M. Wu, Y. Cheng, R. Zhang, W. Han, H. Jiang, C. Bi, Z. Zhang, M. Ye, X. Lin, and Z. Liu, Molecular mechanism and therapeutic strategy of bile acids in Alzheimer’s disease from the emerging perspective of the microbiota-gut-brain axis. Biomedicine & pharmacotherapy = Biomedecine & pharmacotherapie 178 (2024) 117228.

[90] S. MahmoudianDehkordi, M. Arnold, K. Nho, S. Ahmad, W. Jia, G. Xie, G. Louie, A. Kueider-Paisley, M.A. Moseley, J.W. Thompson, L. St John Williams, J.D. Tenenbaum, C. Blach, R. Baillie, X. Han, S. Bhattacharyya, J.B. Toledo, S. Schafferer, S. Klein, T. Koal, S.L. Risacher, M.A. Kling, A. Motsinger-Reif, D.M. Rotroff, J. Jack, T. Hankemeier, D.A. Bennett, P.L. De Jager, J.Q. Trojanowski, L.M. Shaw, M.W. Weiner, P.M. Doraiswamy, C.M. van Duijn, A.J. Saykin, G. Kastenmuller, R. Kaddurah-Daouk, I. Alzheimer’s Disease Neuroimaging, and C. the Alzheimer Disease Metabolomics, Altered bile acid profile associates with cognitive impairment in Alzheimer’s disease-An emerging role for gut microbiome. Alzheimers Dement 15 (2019) 76–92.

[91] J. Xu, S. Patassini, P. Begley, S. Church, H.J. Waldvogel, R.L.M. Faull, R.D. Unwin, and G.J.S. Cooper, Cerebral deficiency of vitamin B5 (d-pantothenic acid; pantothenate) as a potentially-reversible cause of neurodegeneration and dementia in sporadic Alzheimer’s disease. Biochem Biophys Res Commun 527 (2020) 676–681.

[92] M. Scholefield, S.J. Church, J. Xu, S. Patassini, N.M. Hooper, R.D. Unwin, and G.J.S. Cooper, Substantively Lowered Levels of Pantothenic Acid (Vitamin B5) in Several Regions of the Human Brain in Parkinson’s Disease Dementia. Metabolites 11 (2021).

[93] N. Kumari, H. Prentice, and J.Y. Wu, Taurine and its neuroprotective role. Adv Exp Med Biol 775 (2013) 19–27.

[94] P. Singh, K. Gollapalli, S. Mangiola, D. Schranner, M.A. Yusuf, M. Chamoli, S.L. Shi, B. Lopes Bastos, T. Nair, A. Riermeier, E.M. Vayndorf, J.Z. Wu, A. Nilakhe, C.Q. Nguyen, M. Muir, M.G. Kiflezghi, A. Foulger, A. Junker, J. Devine, K. Sharan, S.J. Chinta, S. Rajput, A. Rane, P. Baumert, M. Schonfelder, F. Iavarone, G. di Lorenzo, S. Kumari, A. Gupta, R. Sarkar, C. Khyriem, A.S. Chawla, A. Sharma, N. Sarper, N. Chattopadhyay, B.K. Biswal, C. Settembre, P. Nagarajan, K.L. Targoff, M. Picard, S. Gupta, V. Velagapudi, A.T. Papenfuss, A. Kaya, M.G. Ferreira, B.K. Kennedy, J.K. Andersen, G.J. Lithgow, A.M. Ali, A. Mukhopadhyay, A. Palotie, G. Kastenmuller, M. Kaeberlein, H. Wackerhage, B. Pal, and V.K. Yadav, Taurine deficiency as a driver of aging. Science 380 (2023) eabn9257.

[95] C. Cui, H. Song, Y. Han, H. Yu, H. Li, Y. Yang, and B. Zhang, Gut microbiota-associated taurine metabolism dysregulation in a mouse model of Parkinson’s disease. mSphere 8 (2023) e0043123.

[96] H. Jang, S. Lee, S.L. Choi, H.Y. Kim, S. Baek, and Y. Kim, Taurine Directly Binds to Oligomeric Amyloid-beta and Recovers Cognitive Deficits in Alzheimer Model Mice. Adv Exp Med Biol 975 Pt 1 (2017) 233–241.

[97] S. Ahmed, N. Ma, J. Kawanokuchi, K. Matsuoka, S. Oikawa, H. Kobayashi, Y. Hiraku, and M. Murata, Taurine reduces microglia activation in the brain of aged senescence-accelerated mice by increasing the level of TREM2. Scientific reports 14 (2024) 7427.

