## Supplementary figures and images for "Harmonizing the oral-brain axis: Music-induced microbiota shifts in age-related cognitive disorders and healthy aging"

### Figure S1

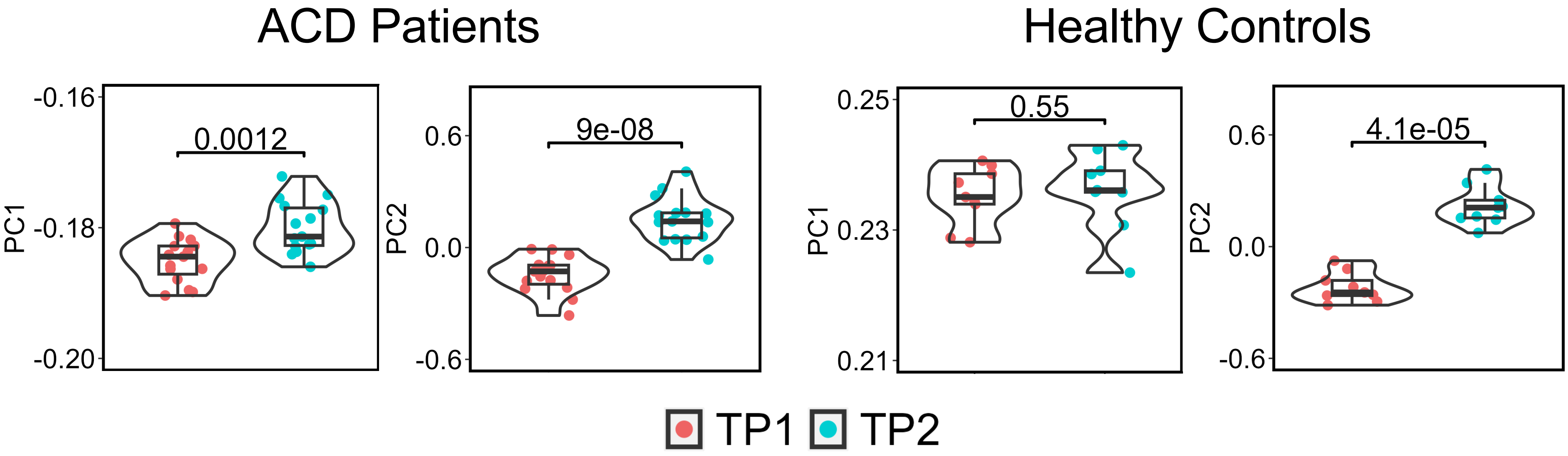

### Figure S2

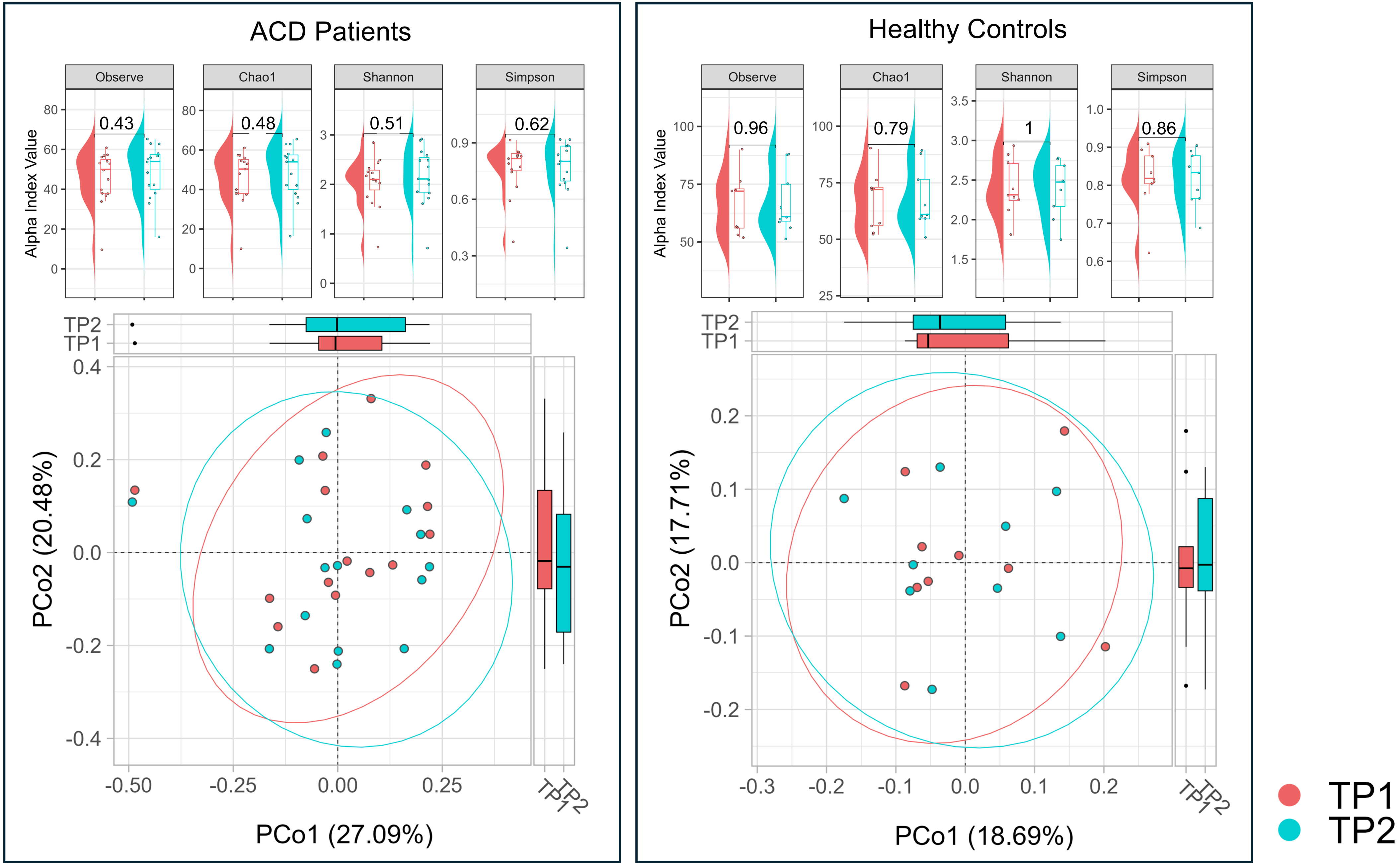

### Figure S3

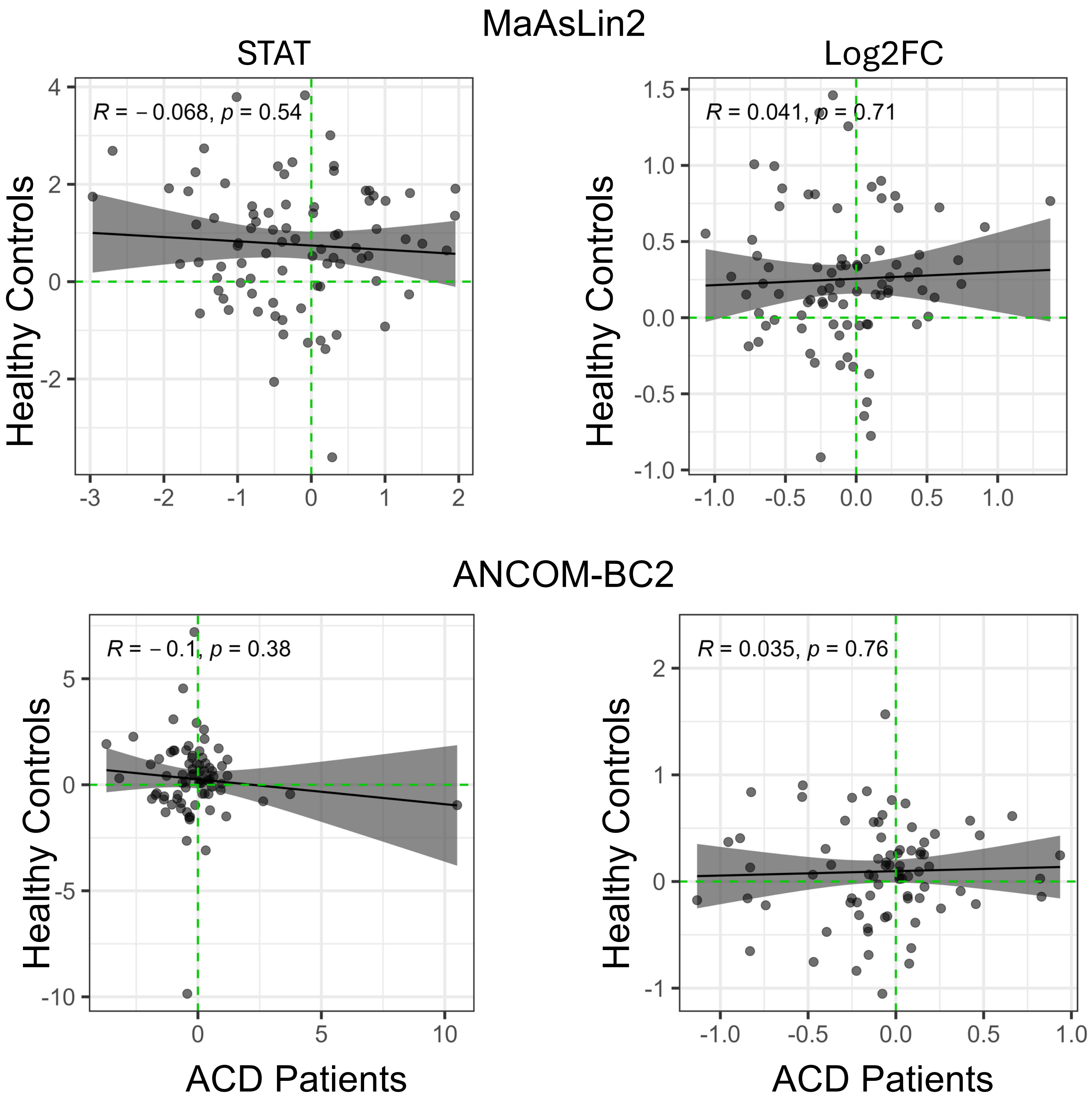
